# Accuracy of automated computer aided-risk scoring systems to estimate the risk of COVID-19 and in-hospital mortality: a retrospective cohort study

**DOI:** 10.1101/2020.12.01.20241828

**Authors:** Muhammad Faisal, Mohammed A Mohammed, Donald Richardson, Massimo Fiori, Kevin Beatson

**Author notes:** Correspondence to Mohammed Amin Mohammed, Faculty of Health Studies, University of Bradford, Richmond Road, BD7 1DP Bradford, UK.

## Abstract

**Objectives:** Although a set of computer-aided risk scoring systems (CARSS), that use the National Early Warning Score and routine blood tests results, have been validated for predicting in-hospital mortality and sepsis in unplanned admission to hospital, little is known about their performance for COVID-19 patients. We compare the performance of CARSS in unplanned admissions with COVID-19 during the first phase of the pandemic.

**Design:** a retrospective cross-sectional study

**Setting:** Two acute hospitals (Scarborough and York) are combined into a single dataset and analysed collectively.

**Participants:** Adult (>=18 years) non-elective admissions discharged between 11-March-2020 to 13-June-2020 with an index NEWS electronically recorded within ±24 hours. We assessed the performance of all four risk score (for sepsis: CARS_N, CARS_NB; for mortality: CARM_N, CARM_NB) according to discrimination (c-statistic) and calibration (graphically) in predicting the risk of COVID-19 and in-hospital mortality.

**Results:** The risk of in-hospital mortality following emergency medical admission was 8.4% (500/6444) and 9.6% (620/6444) had a diagnosis of COVID-19. For predicting COVID-19 admissions, the CARS_N model had the highest discrimination 0.73 (0.71 to 0.75) and calibration slope 0.81 (0.72 to 0.89). For predicting in-hospital mortality, the CARM_NB model had the highest discrimination 0.84 (0.82 to 0.75) and calibration slope 0.89 (0.81 to 0.98).

**Conclusions:** Two of the computer-aided risk scores (CARS_N and CARM_NB) are reasonably accurate for predicting the risk of COVID-19 and in-hospital mortality, respectively. They may be clinically useful as an early warning system at the time of admission especially to triage large numbers of unplanned hospital admissions because they are automated and require no additional data collection.

**Article Summary:**

- In this study, we found that two of the automated computer-aided risk scores are reasonably accurate for predicting the risk of COVID-19 and in-hospital mortality, respectively.
- They may be clinically useful as an early warning system at the time of admission especially to triage large numbers of unplanned hospital admissions because they are automated and require no additional data collection.
- Although we focused on in-hospital mortality (because we aimed to aid clinical decision making in the hospital), the impact of this selection bias needs to be assessed by capturing out-of-hospital mortality by linking death certification data and hospital data.
- We identified COVID-19 based on ICD-10 code ‘U071’ which was determined by COVID-19 swab test results (hospital or community) and clinical judgment and so our findings are constrained by the accuracy of these methods

## Introduction

The novel coronavirus SARS-CoV-2, which was declared as a pandemic on 11-March 2020, produces the newly identified disease ‘COVID-19’ in patients with symptoms (Coronaviridae Study Group of the International Committee on Taxonomy of Viruses[1]) which has challenged health care systems worldwide.

Patients with COVID-19 admitted to a hospital can develop severe disease with life-threatening respiratory and/or multi-organ failure [2,3] with a high risk of mortality. It is recommended that patients at risk of deterioration are referred to as critical care. The appropriate early assessment and management of patients with COVID-19 are important in ensuring high-quality care.

In the UK National Health Service (NHS), the patient’s vital signs are monitored and summarised into a National Early Warning Score (NEWS) or its latest version (NEWS2)[4].

We had developed four automated, computer-aided risk scores to predict the patient’s risk of mortality (CARM_N & CARM_NB) and sepsis (CARS_N & CARS_NB) following emergency medical admission to hospital [5–8]. The _N models use NEWS data and the _NB models incorporate routine blood test results. We refer to this suite of four risk equations as computer-aided risk scoring systems (CARSS).

Although the CARSS is developed and externally validated for predicting in-hospital mortality and sepsis in unplanned admissions to hospital, little is known about its performance in unplanned admissions COVID-19 admissions. In this study, we aimed to compare the performance, in unplanned admissions to a teaching hospital during the first phase of the novel coronavirus SARS CoV-2 (COVID-19) pandemic, in predicting the risk of COVID-19 and in-hospital mortality.

## Methods

### Setting & data

Our cohort of unplanned admissions is from two acute hospitals which are approximately 65 kilometres apart in the Yorkshire & Humberside region of England – Scarborough hospital (n∼300 beds) and York Hospital (YH) (n∼700 beds), managed by York Teaching Hospitals NHS Foundation Trust. For this study, the two acute hospitals are combined into a single dataset and analysed collectively. The hospitals have electronic NEWS scores and vital signs recording which is routinely collected as part of the patient’s process of care (see Table S1).

We considered all adult (age≥18 years) emergency medical admissions (excluding ambulatory care area patients), discharged during 3 months (11 March 2020 to 13 June 2020), with electronic NEWS recorded within ±24 hours of admission. This on-admission NEWS score is referred to as the index NEWS.

For each emergency admission, we obtained a pseudonymised patient identifier, patient’s age (years), gender (male/female), discharge status (alive/dead), admission and discharge date and time, diagnoses codes based on the 10th revision of the International Statistical Classification of Diseases (ICD-10) [9] [10], NEWS (including its subcomponents respiratory rate, temperature, systolic pressure, pulse rate, oxygen saturation, oxygen supplementation, oxygen scales 1 & 2, and alertness including confusion) [4,11], blood test results (albumin, creatinine, haemoglobin, potassium, sodium, urea, and white cell count), and Acute Kidney Injury (AKI) score.

We had developed and externally validated four risk scores: 1) CARM_N for predicting in-hospital mortality based on NEWS [8]; 2) CARM_NB for predicting in-hospital mortality that incorporates routine blood test results [5]; CARS_N for predicting sepsis based on NEWS [7]; CARS_NB for predicting sepsis that incorporates routine blood test results [6] (see Table 1). These four equations are collective known as computer-aided risk scoring systems (CARSS), calculated using index NEWS and blood test results. We excluded records where the index NEWS (or blood test results) was not within ±24 hours (±96 hours) or was missing/not recorded at all (see Table S2). The ICD-10 code ‘U071’ was used to identify records with COVID-19. We searched primary and secondary ICD-10 codes for ‘U071’ for identifying COVID-19.

**Table 1:** Four risk scores for predicting the risk of mortality and sepsis, known as computer-aided risk scoring systems (CARSS)

| Computer-Aided Risk (CAR) score | NEWS data only (N) | NEWS and Blood test results data (NB) |
| --- | --- | --- |
| Mortality (M) | CARM_N | CARM_NB |
| Sepsis (S) | CARS_N | CARS_NB |

### Statistical Analyses

We report discrimination and calibration statistics as performance measures for CARSS [12].

We determined the discrimination of CARSS using the concordance statistic (c-statistic) that gives the probability of randomly selected patients who experienced adverse outcome had a higher risk score than a patient who does not. For a binary outcome (alive/died or COIVD-19/Non-Covid-19), the c-statistic is the area under the Receiver Operating Characteristics (ROC) curve [13]. The ROC curve is a plot of the sensitivity, (true positive rate), versus 1-specificity, (false positive rate), for consecutive predicted risks. A c-statistic of 0.5 is no better than tossing a coin, whilst a perfect model has a c-statistic of 1. In general, values less than 0.7 are considered to show poor discrimination, values of 0.7 to 0.8 can be described as reasonable, and values above 0.8 suggest good discrimination[14].

Calibration measures a model’s ability to generate predictions that are on average close to the average observed outcome and can be readily seen on a scatter plot (y-axis observed risk, x-axis predicted risk). Perfect predictions should be on the 45° line. We internally validated and assessed the calibration for all the models using the bootstrapping approach [15,16]. The overall statistical performance was assessed using the scaled Brier score which incorporates both discrimination and calibration [12]. The Brier score is the squared difference between actual outcomes and predicted risk of death, scaled by the maximum Brier score such that the scaled Brier score ranges from 0– 100%. Higher values indicate superior models.

We followed the STROBE guidelines to report the findings [17]. All analyses were undertaken using R [18] and Stata [19].

### Ethical Approval

This study used de-identified data and received ethical approval from the Health Research Authority (HRA) and Health and Care Research Wales (HCRW) (reference number 19/HRA/0548).

### Patient and Public Involvement

No patient involved

## Results

### Cohort description

There were 6480 discharges over 3 months. We excluded 36 (0.6%) records because the index NEWS was not recorded within ±24 hours of the admission date/time or NEWS was missing or not recorded at all (see Table S2). We further excluded 1175 (18.1%) because no or missing blood test results recorded.

The prevalence of COVID-19 was 9.6% (620/6444) and of these 32% (199/620) discharged deceased. The demographic, vital signs and outcome profiles of the COVID-19 versus non-COVID-19 admissions and discharged deceased versus discharged alive are shown in Table 2 and Figure S1-S4. COVID-19 admissions were older (73.3 vs 67.7, p<0.001), more likely to be male (54.7% vs 50.1%, p<0.001), with higher index NEWS (4.0 vs 2.5, p<0.001). They also had longer hospital stay (7.3 days vs 3.0 days, p<0.001) and higher in-hospital mortality (32.1% vs 5.8%, p<0.001). The average CARSs (CARM_N, CARM_NB, CARS_N, CARS_NB) risk was higher for COVID-19 admissions and for those who discharged deceased.

**Table 2:** Characteristics of emergency medical admissions in COVID-19 versus non-COVID-19 who discharged alive/deceased.

| Characteristic | COVID-19 |  | Non-COVID-19 |  |
| --- | --- | --- | --- | --- |
|  | Discharged<br>Deceased | Discharged<br>Alive | Discharged<br>Deceased | Discharged<br>Alive |
| N | 199 | 421 | 336 | 5488 |
| Median Length of Stay (IQR) | 9.61 (14.43) | 6.73 (10.52) | 4.72 (8.88) | 2.96 (5.28) |
| Male (%) | 123 (61.81) | 216 (51.31) | 169 (50.3) | 2749 (50.09) |
| Mean Age [years] (SD) | 80.22 (10.01) | 70.08 (16.43) | 79.44 (12.65) | 67.02 (19.14) |
| Mean NEWS (SD) | 4.94 (3.02) | 3.52 (2.5) | 4.89 (3.42) | 2.33 (2.08) |
| Mean CARM_N (SD) | 0.14 (0.11) | 0.06 (0.07) | 0.15 (0.13) | 0.04 (0.06) |
| Mean CARM_NB (SD) | 0.15 (0.15) | 0.06 (0.08) | 0.16 (0.17) | 0.04 (0.06) |
| Mean CARS_N (SD) | 0.36 (0.19) | 0.25 (0.16) | 0.28 (0.16) | 0.16 (0.13) |
| Mean CARS_NB (SD) | 0.34 (0.2) | 0.21 (0.16) | 0.29 (0.18) | 0.15 (0.13) |

*\* Blood test results are missing 1175 (18.1%)*

We assessed the four CARSs models (CARM_N, CARM_NB, CARS_N, CARS_NB) performance according to discrimination (c-statistic) and calibration (graphically) in predicting the risk of COVID-19 and in-hospital mortality (see Table 3 and Figure 2 & 3).

**Table 3:** Performance of CARSs models for predicting the risk of COVID-19 and in-hospital mortality.

| Outcome | Model | Mean risk without adverse outcome | Mean risk with adverse outcome | Absolute risk difference | Scaled brier score | Discrimination AUC (95% CI) | Calibration Slope (95% CI) |
| --- | --- | --- | --- | --- | --- | --- | --- |
| COVID-19 | CARM_N | 0.09 | 0.15 | 0.06 | -0.02 | 0.68<br>(0.66 to 0.70) | 0.47<br>(0.41 to 0.54) |
| COVID-19 | CARM_NB | 0.09 | 0.17 | 0.08 | -0.05 | 0.68<br>(0.65 to 0.70) | 0.37<br>(0.31 to 0.43) |
| COVID-19 | CARS_N | 0.09 | 0.17 | 0.08 | 0.05 | 0.73<br>(0.71 to 0.75) | 0.81<br>(0.72 to 0.89) |
| COVID-19 | CARS_NB | 0.09 | 0.16 | 0.07 | 0.01 | 0.68<br>(0.66 to 0.70) | 0.56<br>(0.47 to 0.64) |
| In-hospital Mortality | CARM_N | 0.07 | 0.21 | 0.14 | 0.14 | 0.82<br>(0.81 to 0.84) | 1.06<br>(0.96 to 1.14) |
| In-hospital Mortality | CARM_NB | 0.07 | 0.25 | 0.18 | 0.15 | 0.84<br>(0.82 to 0.85) | 0.89<br>(0.81 to 0.98) |
| In-hospital Mortality | CARS_N | 0.08 | 0.16 | 0.08 | 0.07 | 0.77<br>(0.75 to 0.79) | 0.95<br>(0.85 to 1.04) |
| In-hospital Mortality | CARS_NB | 0.08 | 0.18 | 0.10 | 0.08 | 0.78<br>(0.76 to 0.80) | 0.92<br>(0.82 to 1.01) |

**Figure 1.**
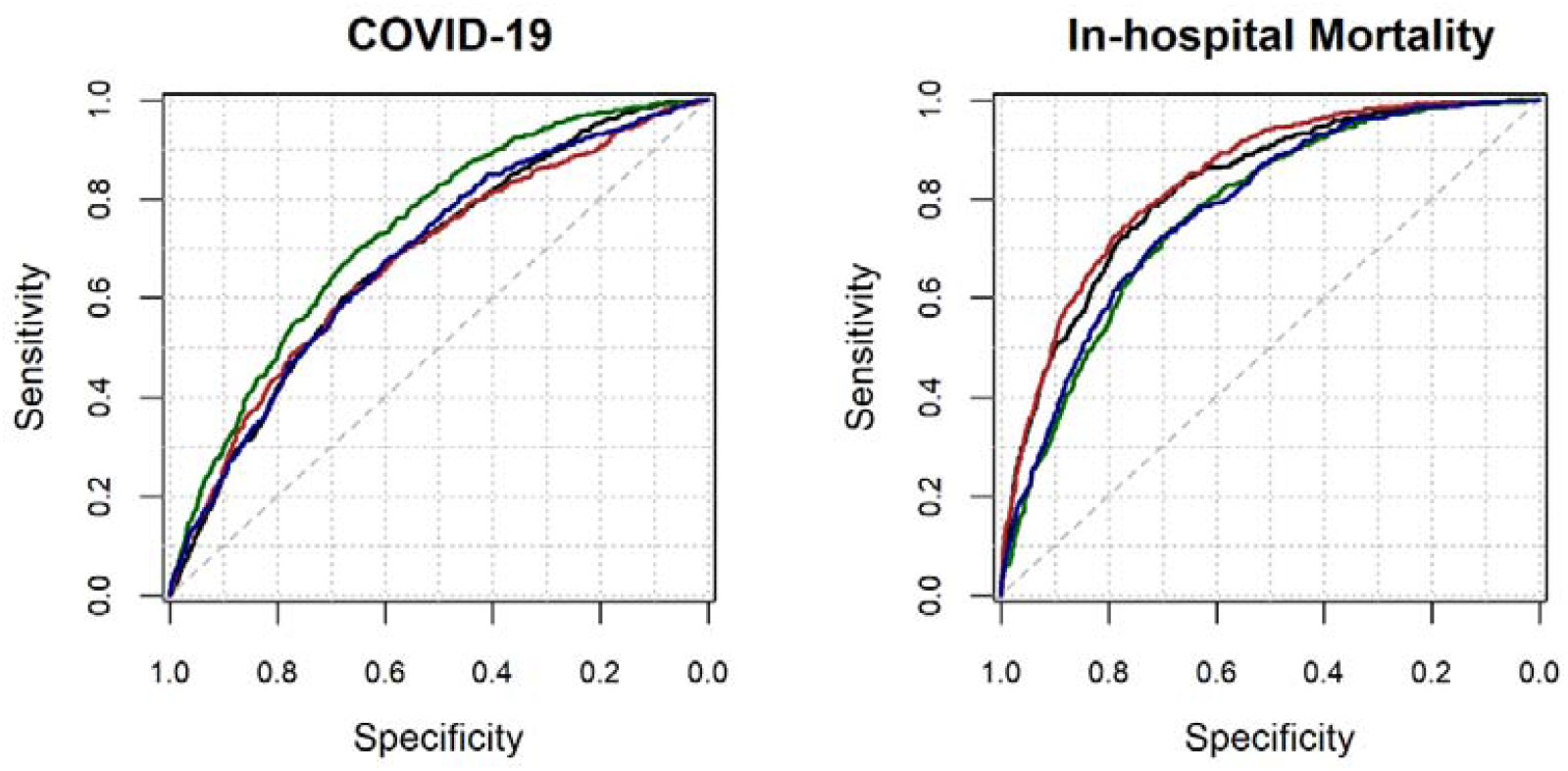
Receiver Operating Characteristic curve for four CARSS models in predicting the risk of COVID-19 (left) and in-hospital mortality (right) The black line is for CARM_N and the red line is for CARM_NB; The green line is CARS_N and the blue line is for CARS_NB

**Figure 2.**
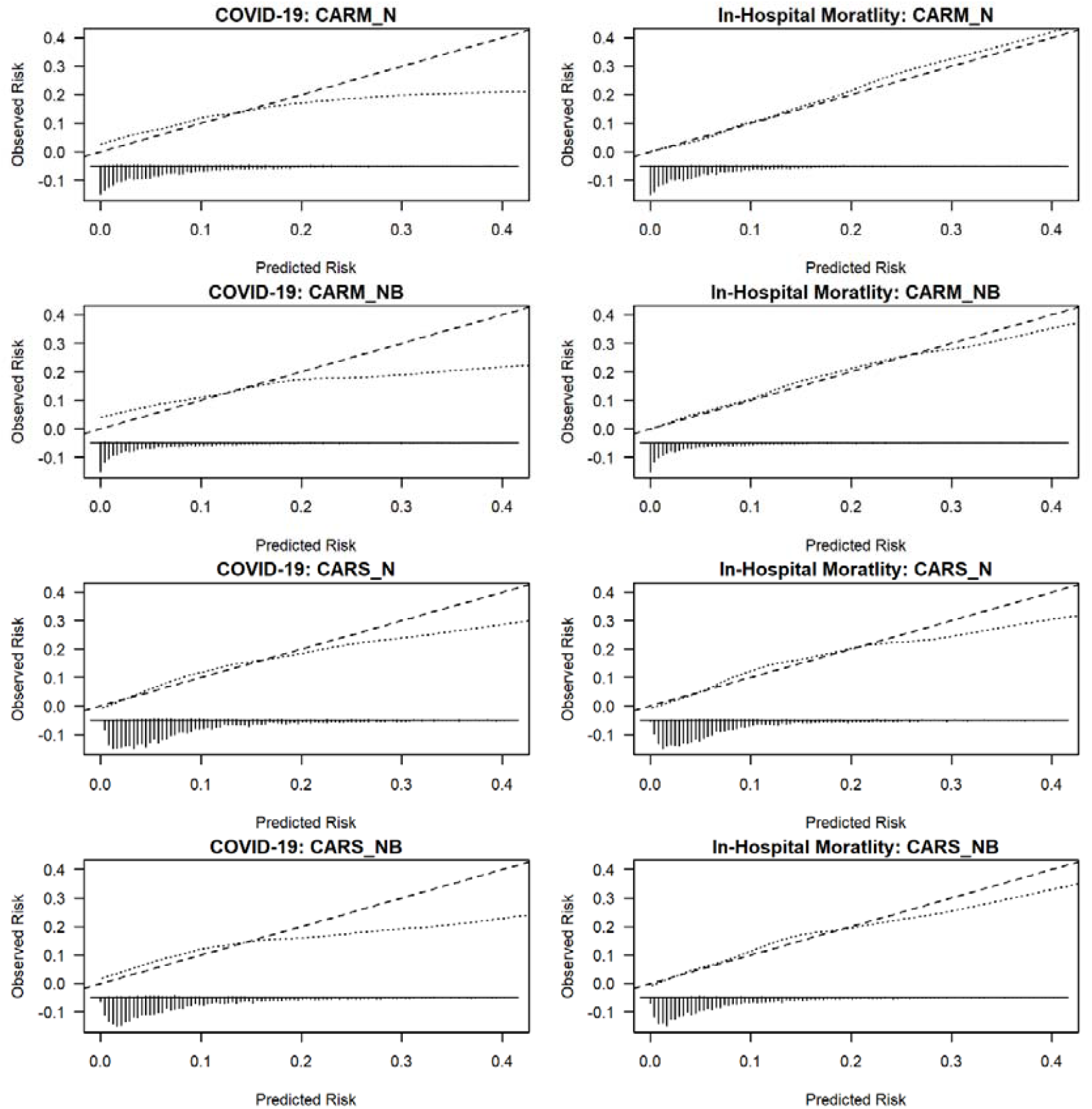
External validation of CARSS models, respectively for predicting the risk of COVID-19 (left column) and in-hospital mortality (right column) NB: We limit the risk of mortality to 0.40 for visualisation purpose because beyond this point, we have few patients.

For predicting COVID-19 admissions, the CARS_N model performed better than others in terms of discrimination 0.73 (0.71 to 0.75) and calibration slope 0.81 (0.72 to 0.89).

For predicting in-hospital mortality, the CARM_NB model performed better than others in terms of discrimination 0.84 (0.82 to 0.75) and calibration slope 0.89 (0.81 to 0.98).

## Discussion

We assessed the performance of four computer aided risk scores and found that the CARS_N (which predicts sepsis based on NEWS) and CARM_NB (which predicts mortality based on NEWS and blood test results) accurately predict the risk of COVID-19 and in-hospital mortality, respectively.

Not surprisingly, CARM_NB is found to be good discrimination and calibration which is consistent with the literature [5,20] as CARM_NB was developed for predicting the in-hospital mortality. However, CARS_N was developed for predicting sepsis and we found it has good discrimination and calibration compared to other CARSS models. Interestingly, several features are common in COVID-19 and sepsis patients [21]. Zhou et al. [21] found that the Sequential Organ Failure Assessment (SOFA) score (for sepsis) is associated with in-hospital mortality in COVID-19 patients. Although further studies are need to investigate the common features of sepsis and COVID-19, it is clear that hyper inflammation and coagulopathy contribute to disease severity and death in COVID-19 patients [22]. A recent systematic review identified models to predict mortality from COVID-19 with c-statistics that ranged from 0.87 to 1 [23]. However, despite these high c-statistics, the review authors cautioned against the use of these models in clinical practice because of the high risk of bias and poor reporting of studies which are likely to have led to optimistic results [23].

The main advantages of our models are that they are designed to incorporate data which are already available in the patient’s electronic health record thus placing no additional data collection or computational burden on clinicians and are readily automated.

There are several limitations to our study. (1) This is data from a single NHS Trust and the extent to which these findings are generalisable, further study is required. (2) We used the index NEWS and blood test results which reflects the ‘on-admission’ risk of mortality of the patients. Nonetheless, NEWS and blood test results are repeatedly updated for each patient according to local hospital protocols (Figure S5 in supplementary material). (3) Although we focused on in-hospital mortality (because we aimed to aid clinical decision making in the hospital), the impact of this selection bias needs to be assessed by capturing out-of-hospital mortality by linking death certification data and hospital data. (4) We identified COVID-19 based on ICD-10 code ‘U071’ which was determined by COVID-19 swab test results (hospital or community) and clinical judgment and so our findings are constrained by the accuracy of these methods [24,25].

## Conclusion

The CARS_N and CARM_NB are reasonably accurate for predicting the risk of COVID-19 and in-hospital mortality, respectively. They may be clinically useful for an early warning system at the time of admission especially to triage large numbers of unplanned hospital admissions.

## Data Availability

Our data sharing agreement with NHS York hospital trust does not permit us to share this data with other parties. Nonetheless, if anyone is interested in the data, then they should contact the R&D offices in the first instance.

## Contributorship

DR & MAM had the original idea for this work. MF undertook the statistical analyses with guidance from MAM. MFi & KB extracted the necessary data frames. DR gave a clinical perspective. DR, MF and MAM wrote the first draft of this paper and all authors subsequently assisted in redrafting and have approved the final version.

