## supplementary material for "Accuracy of automated computer aided-risk scoring systems to estimate the risk of COVID-19 and in-hospital mortality: a retrospective cohort study"

| **Physiological Parameters** | **3** | **2** | **1** | **0** | **1** | **2** | **3** |
| --- | --- | --- | --- | --- | --- | --- | --- |
| **Respiration Rate** | **≤8** |  | **9 - 11** | **12 - 20** |  | **21 - 24** | **≥25** |
| **Oxygen Saturations** | **≤91** | **92 - 93** | **94 - 95** | **≥96** |  |  |  |
| **Any Supplemental Oxygen** |  | **Yes** |  | **No** |  |  |  |
| **Temperature** | **≤35.0** |  | **35.1 - 36.0** | **36.1 - 38.0** | **38.1 - 39.0** | **≥39.1** |  |
| **Systolic BP** | **≤90** | **91 - 100** | **101 - 110** | **111 - 219** |  |  | **≥220** |
| **Heart Rate** | **≤40** |  | **41 - 50** | **51-90** | **91 - 110** | **111 - 130** | **≥131** |
| **Level of Consciousness** |  |  |  | **Alert** |  |  | **Voice, Pain, or Unconscious** |

**Table S1: NEWS scoring chart**

The NEWS [https://www.rcplondon.ac.uk/projects/outputs/national-early-warning-score-news] is based on a scoring system in which a score is allocated to vital signs physiological measurements already undertaken when patients present to or are being monitored in hospital. A score is allocated to each as they are measured, the magnitude of the score reflecting how extreme the parameter varies from the norm. This score is then aggregated and uplifted for people requiring oxygen.

| **Characteristic** | **COVID-19** | **Non-COVID-19** | **All** |
| --- | --- | --- | --- |
|  | N (%) | N (%) | N (%) |
| **Total emergency medical discharges between**  **11 Mar 20 to 13 June 20** | 622 | 5858 | 6480 |
| **Excluded: No NEWS recorded (%)** | 0 (0.0) | 19 (0.3) | 19 (0.3) |
| **Excluded: First NEWS after 24 hours of admission (%)** | 2 (0.3) | 15 (0.3) | 17 (0.3) |
| **Excluded: No or missing blood test results recorded (%)** | 111 (17.8) | 1064 (18.1) | 1175 (18.1) |
| **Total excluded (%)** | 113 (18.2) | 1098 (18.9) | 1211 (18.7) |
| **Total included (%)** | 509 (81.8) | 4760 (81.1) | 5269 (81.3) |

**Table S2 Number of emergency medical admissions included/excluded**

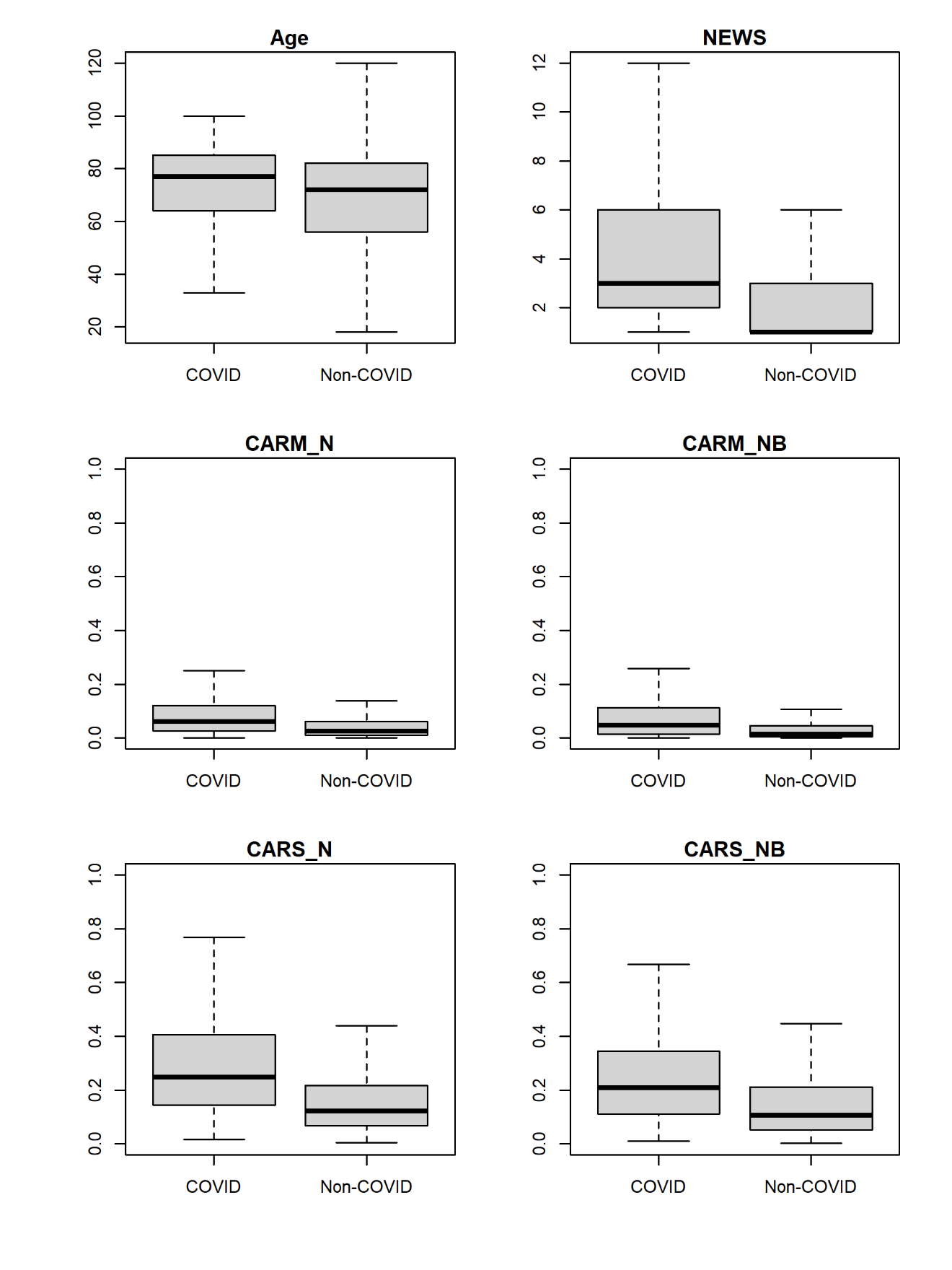

### Figure S1 Boxplot for age, NEWS and CARSS without outliers with respect to COVID-19 status (COVID/Non-COVID)

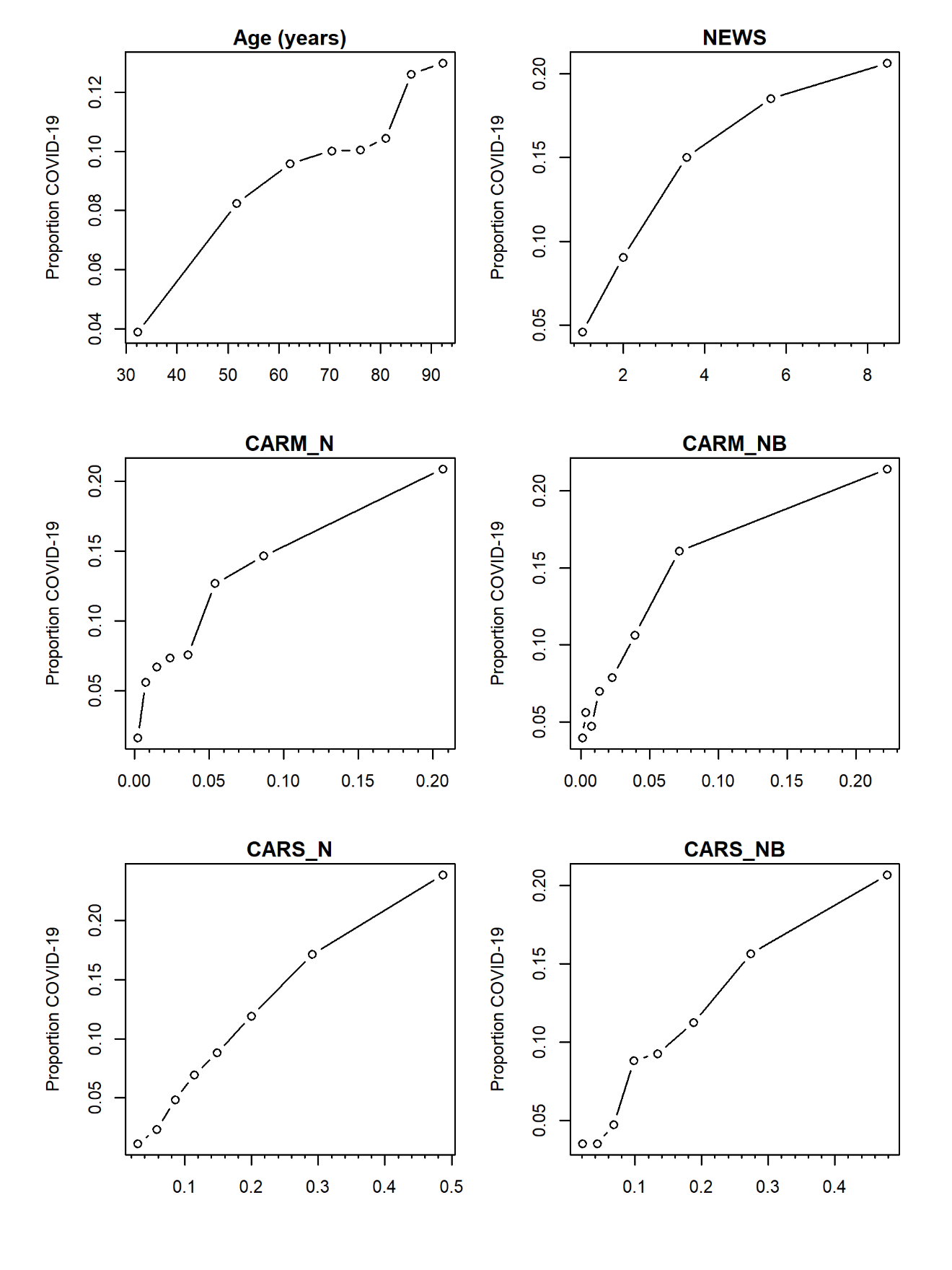

Figure S2 Scatter plots showing the observed risk of COVID-19 with age, NEWS, and CARSS

NB: y-axis range changes in each plot.

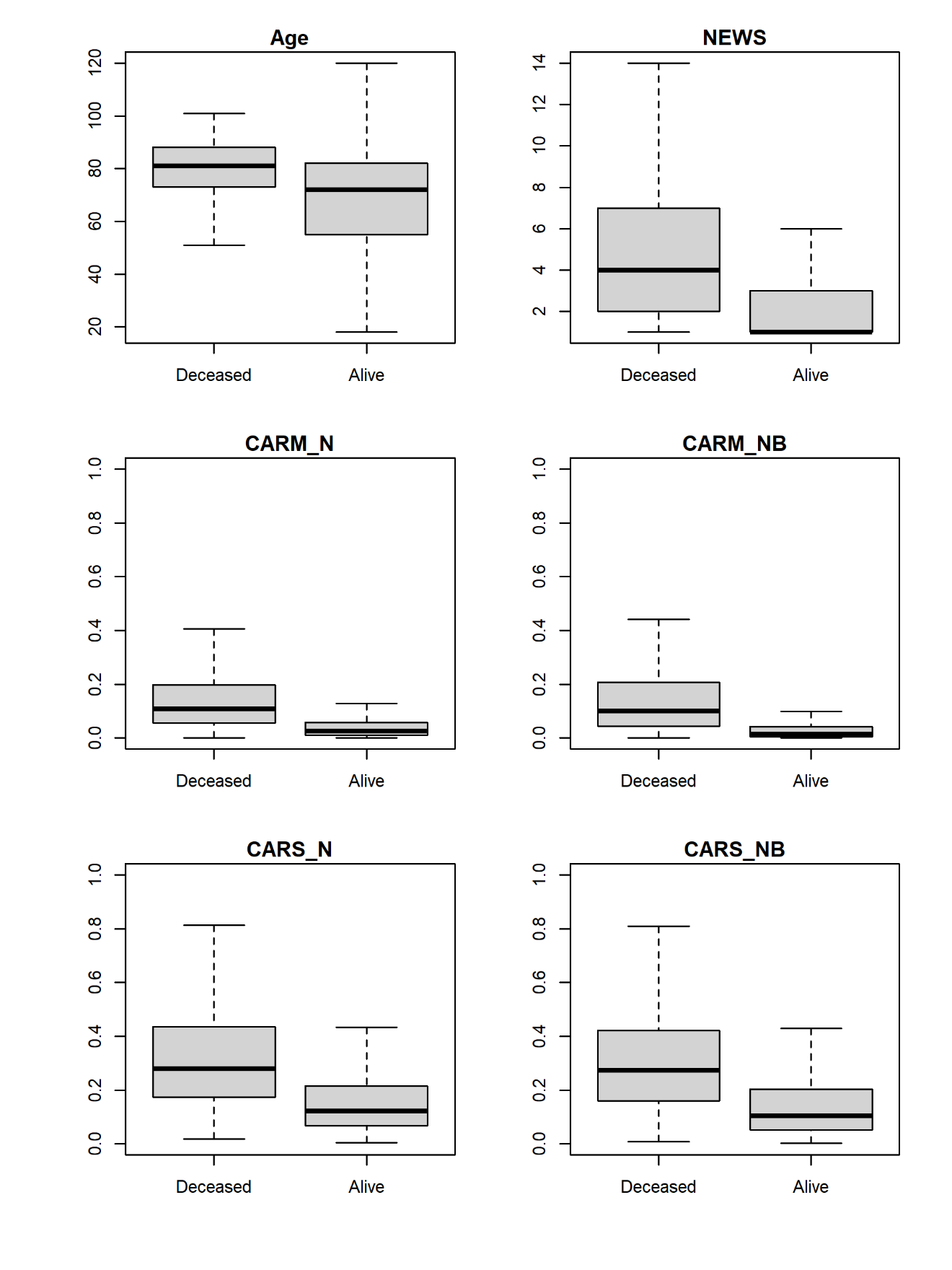

Figure S3 Boxplot for age, NEWS, and CARSS without outliers with respect to discharged status (Deceased/Alive).

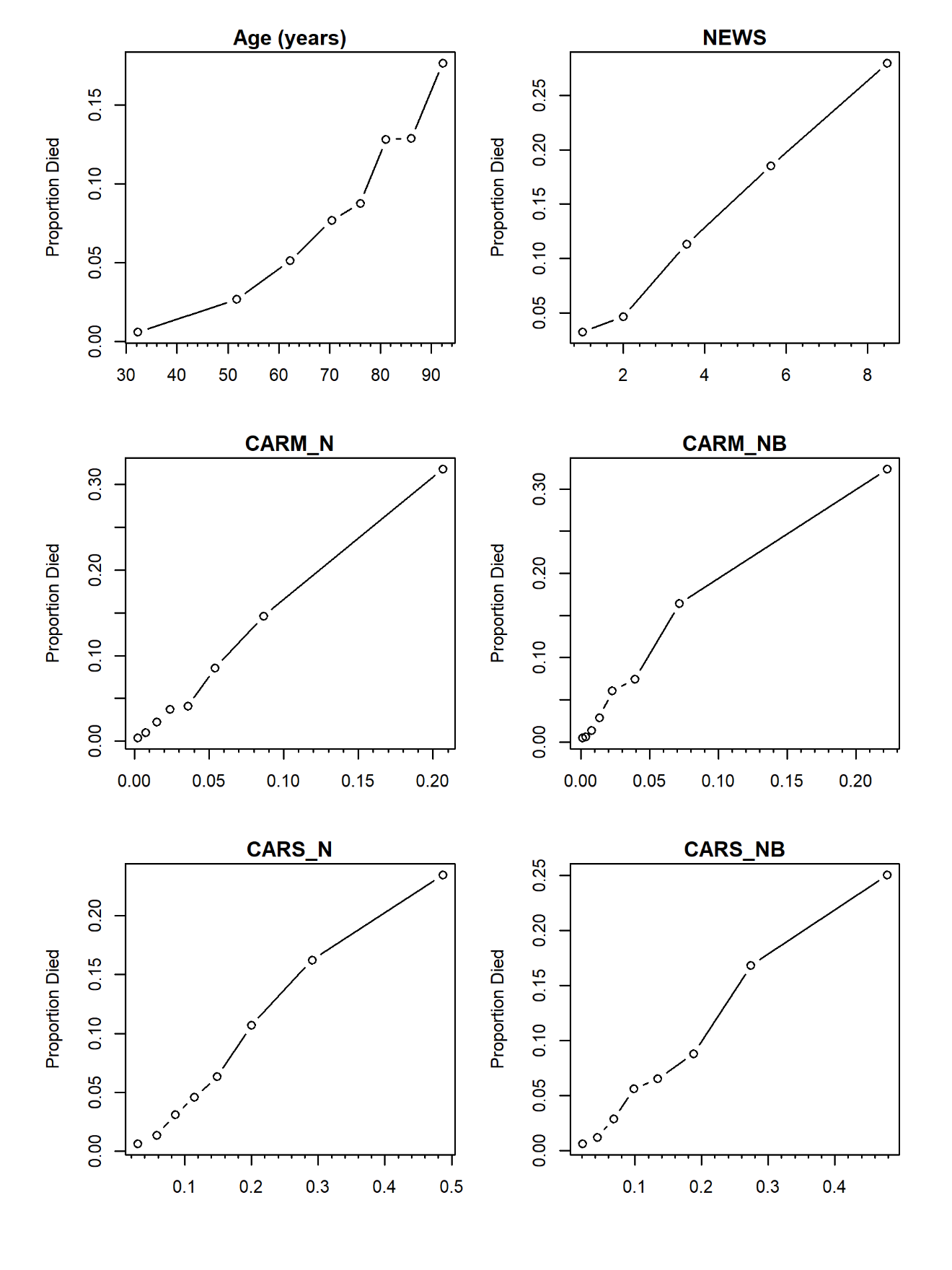

Figure S4 Scatter plots showing the observed risk of in-hospital mortality with age, NEWS, and CARSS

NB: y-axis range changes in each plot.

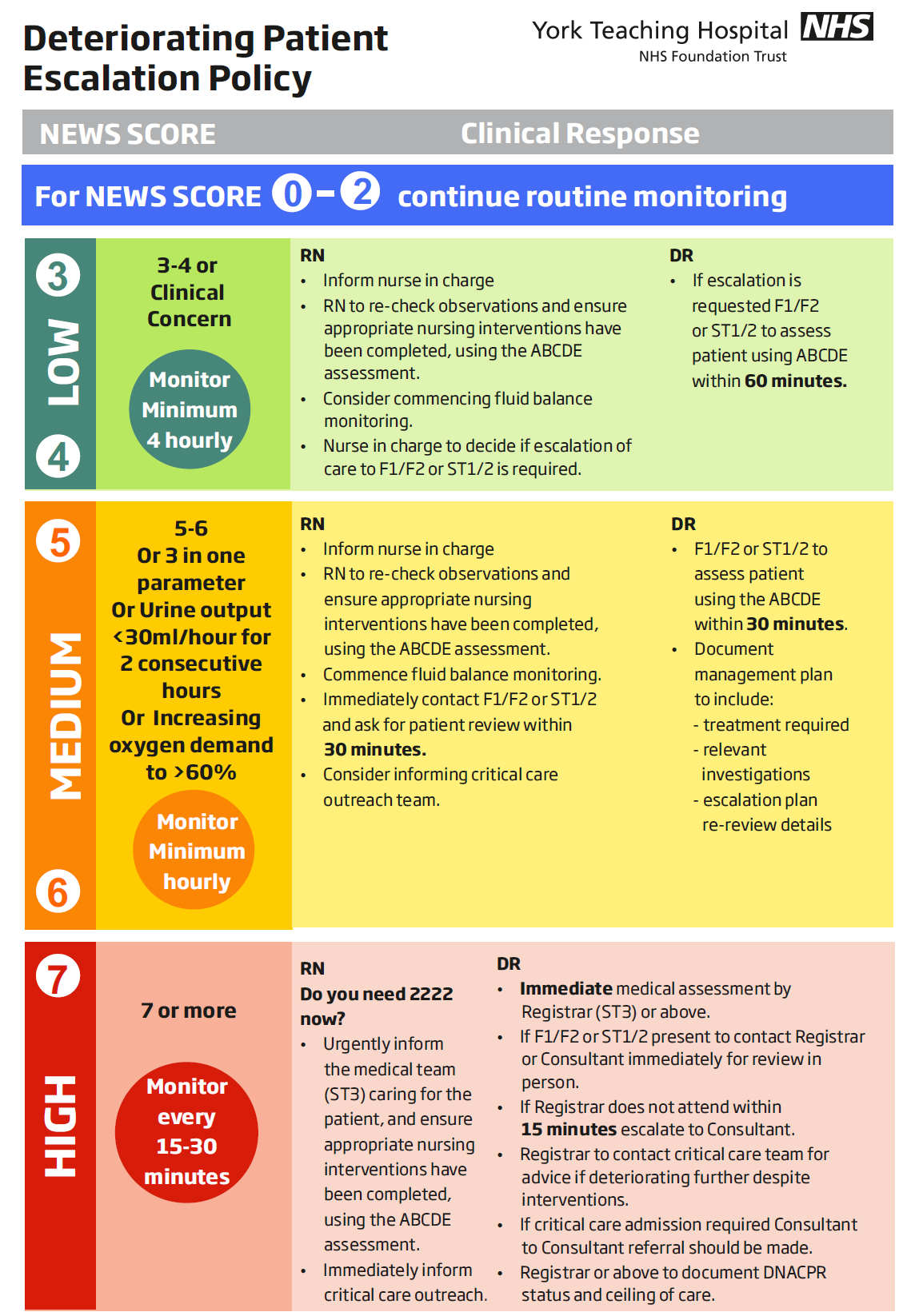

**Figure S5 Escalation policy of deteriorating patients in York Teaching Hospital NHS Foundation Trust**
